# Insufficient Water, Sanitation, and Healthcare Facilities Hinder Schistosomiasis Control in Endemic Areas: A Mixed Methods Study

**DOI:** 10.1101/2024.08.30.24312829

**Authors:** Aspire Mudavanhu, Rachelle Weeda, Maxson Kenneth Anyolitho, Linda Mlangeni, Luc Brendonck, Tawanda Manyangadze, Tine Huyse

## Abstract

**Introduction:** Schistosomiasis remains a significant public health issue in sub-Saharan Africa, particularly in resource-limited settings. This mixed-methods study assesses the knowledge, attitudes, and practices (KAP) related to schistosomiasis in two communities in Zimbabwe’s Chiredzi district: Hippo Valley Estate (HVE) and Chipimbi.

**Methodology:** In August 2022, a total of 279 household adults were surveyed using semi-structured questionnaires, selected through systematic random sampling. Additionally, four key informants were interviewed, and eight focus group discussions (FGDs) were conducted. Cramer’s V (φ) and Gamma (γ) coefficients were used to determine associations between sociodemographic factors and KAP variables, with a p-value of 0.05 indicating statistical significance. Quantitative data were analyzed using frequencies, percentages, and chi-square tests, while qualitative data were analyzed by identifying themes and sub-themes.

**Results:** Awareness of schistosomiasis was high, with 87.5% of respondents having heard of the disease and 86% knowing its transmission modes and symptoms. However, misconceptions persist, such as the belief that walking barefoot or eating unwashed fruits are major risk factors. Only 7% recognized the importance of avoiding unsafe water, a challenge further amplified in both communities due to their reliance on water for irrigation in HVE and as a primary water source in Chipimbi, especially when stored water is depleted. Although 98% emphasized the importance of latrine use, open defecation remains prevalent due to insufficient latrine coverage in Chipimbi (14%) and occasional water shortages for flushing in HVE. Both communities showed positive attitudes toward treatment, but access to healthcare facilities remains a significant barrier due to selective treatment, long distances, and inadequate facilities.

**Conclusion:** Despite high awareness and positive attitudes, inadequate water, sanitation, and healthcare facilities hinder effective schistosomiasis control. Enhancing community-based awareness, improving access to clean water, and increasing latrine coverage are crucial steps toward sustainable schistosomiasis management

**Author Summary:** Schistosomiasis, a neglected tropical disease, remains a persistent public health challenge, particularly in resource-limited areas. In Zimbabwe, despite six rounds of mass drug administration, with a general prevalence rate of 23%. This study examines two contrasting communities in Zimbabwe’s Chiredzi district: Hippo Valley Estate (HVE), which has benefited from extensive interventions including safe water provision, high latrine coverage, snail control, and regular treatment programs, and Chipimbi, which has not received any such interventions. Both communities are located in an area heavily impacted by large-scale sugarcane irrigation, a factor known to exacerbate schistosomiasis transmission. The study assesses the knowledge, attitudes, and practices (KAP) related to schistosomiasis through surveys and focus group discussions. Findings reveal high awareness of schistosomiasis and its symptoms in both communities, but with persistent misconceptions about risk factors. Despite positive attitudes toward treatment, significant barriers remain, particularly in access to healthcare. Moreover, risky practices like open defecation and unsafe water contact persist due to inadequate water and sanitation infrastructure. This study underscores the need for continued and tailored public health interventions that address misconceptions, improve infrastructure, and enhance healthcare access to effectively control schistosomiasis in these communities.

## Introduction

Schistosomiasis, commonly known as bilharzia, is a waterborne parasitic disease categorized as a neglected tropical disease (NTD). NTDs historically receive less attention from the research community, disproportionately impacting impoverished communities worldwide [1]. Endemic in 78 countries, schistosomiasis puts approximately 779 million people at risk [2], with over 230 million estimated infections globally as of 2014 [3]. Sub-Saharan Africa, home to two-thirds of the global extremely poor population, bears over 90% of schistosomiasis cases [2]. Considered a disease of poverty, schistosomiasis particularly affects those living in poor conditions, with limited access to safe water and proper sanitation [4].

The parasites responsible for schistosomiasis belong to the genus *Schistosoma* (Trematoda: Schistosomatidae), with a complex, multi-host lifecycle involving a freshwater snail and a human host. Two main forms of human schistosomiasis exist i.e. urinary schistosomiasis caused by *Schistosoma haematobium*, characterized by pain during urination and hematuria [3]; and intestinal schistosomiasis caused by *Schistosoma mansoni*, characterized by symptoms like abdominal pain, blood in stool, and liver enlargement [5]. Untreated infections can lead to severe health complications, including infertility, liver fibrosis, anemia, and impaired cognition [3]. Due to these lifelong impacts, schistosomiasis not only threatens individual and household health but also imposes significant social and financial burdens [6,7].

Our study took place in Zimbabwe, a country where schistosomiasis is endemic, affecting 91% of its districts and presenting a nationwide prevalence of 22.7% [8]. Following WHO 2012 recommendations, Zimbabwe so far implemented at least six rounds of mass drug administration (MDA) targeting school-aged children, resulting in a significant reduction in the intensity and morbidity prevalence of schistosomiasis in recipient communities [9]. Over the past two decades, this strategy has contributed to a nearly 60% reduction in schistosomiasis prevalence among school-aged children across sub-Saharan Africa [10]. However, MDA’s reliance has limitations, as it does not prevent reinfection, and the untreated population continues to contaminate the environment through ongoing water contact [11]. Addressing socio-ecological factors becomes imperative for designing effective control programs [4,12]. To achieve sustainable prevention, considerations should include snail control, improved treatment coverage, enhanced water, sanitation, and hygiene (WASH) interventions, along with behavioral change interventions [2,13].

Schistosomiasis, being a complex disease intertwined with environmental and socio-economic aspects, poses unique challenges, especially in a culturally diverse nation like Zimbabwe. Factors such as ethnicity, religion, gender, and education levels but also WASH facilities and interventions can significantly shape the knowledge, attitudes, and practices (KAP) of communities regarding schistosomiasis [14] yet a dearth of studies exploring this crucial component exists. Recognizing this complexity, context-specific research at micro-geographical levels becomes crucial to understanding the socio-ecological factors influencing schistosomiasis infection and affecting the success of control interventions [15]. To that effect, we conducted a comprehensive community-based survey, utilizing both quantitative and qualitative approaches, focusing on the Chiredzi district of Zimbabwe. We selected two main study areas that differ in socio-economic aspects such as access to WASH infrastructure and past control interventions. This study aimed to provide valuable insights into the local knowledge, attitudes, and practices surrounding schistosomiasis, and to test for a possible influence of these socio-economic differences. The overall aim is to contribute to the optimization of control programs, particularly in an area identified as one of the country’s most severely affected by schistosomiasis.

## Methodology

### Research design

We employed a mixed methods approach with an equal status design to evaluate the existing levels of knowledge, attitudes, and practices of individuals in the study area regarding schistosomiasis. This comprehensive strategy involved the collection and analysis of both qualitative and quantitative data, recognizing their complementary nature [16]. Quantitative data were acquired through pre-structured questionnaires, while qualitative insights were obtained through focus group discussions (FGDs) and key informant interviews (KIIs). This dual-method approach aimed to provide a more holistic understanding of the subject.

### Study area and setting

In August 2022, we conducted a cross-sectional study in Chiredzi, a district in Zimbabwe with reported high prevalence rates of *Schistosoma mansoni* (43.7%) and *Schistosoma haematobium* (43.7%) [8]. Our focus was on the communities of Hippo Valley Estate (HVE) and Chipimbi (Fig. 1), which have contrasting access to water, sanitation, and healthcare facilities.

**Fig 1.**
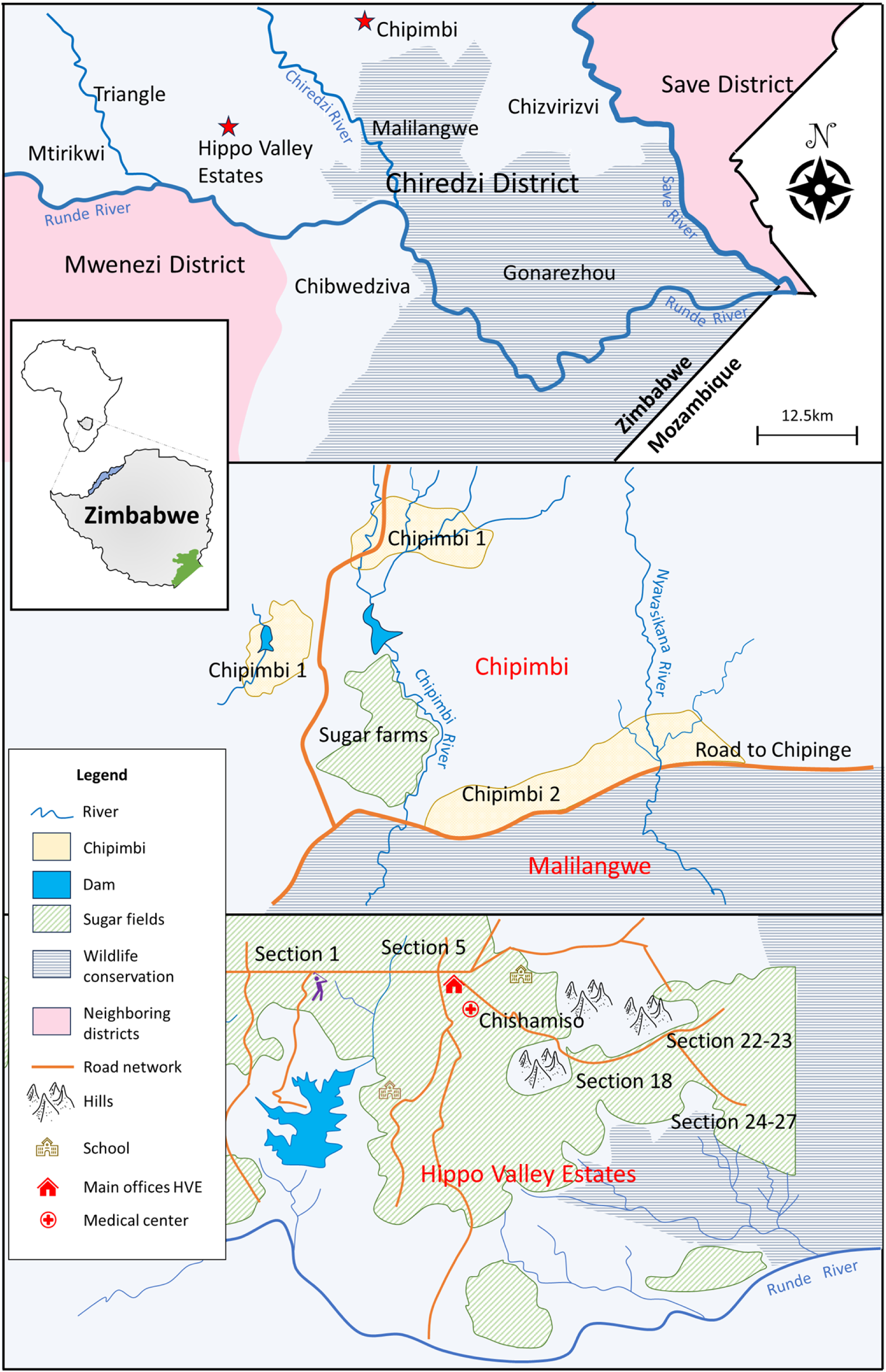
Map of the study area depicting villages and sections from which study participants were recruited. (A) Position of the study area in Zimbabwe (shaded black) on the map of Africa and the location of the study region (Chiredzi, shown as in green on the south-eastern part) on the map of Zimbabwe. (B) Map of Chiredzi showing various communities of the district. (C) Map of Chipimbi Community further divided in two: Chipimbi 1 comprise of villages 1 to 5 whereas Chipimbi 2 comprises of villages 6 to 10. (D) Map of Hippo Valley Estates showing the various sections and how they morph into areas based on the organization’s internal disease surveillance plan.

Villages in HVE, referred to here as sections, each have a satellite clinic in addition to the main medical center. Managed by Tongaat Hulett, a multinational company, HVE has a long history of disease control programs dating back to 1971 [17]. These programs include snail control, an annual chemotherapy component targeting school children, and an intensive water and sanitation component. Assessments of these control programs [18,19] have shown a significant decline in both prevalence and intensity of infections over long periods, with a sustained phase of prevalence below 10%. However, a recent malacological survey indicated that the intermediate host snails, *Biomphalaria* and *Bulinus* spp., still persist in HVE in high numbers – 60 and 122, respectively – although they were not infected [20]. Given the irrigation practices in this region, it is reasonable to anticipate that many people are still exposed to infection.

In contrast, the clinic in Chipimbi remains unused, forcing residents to travel to the main district clinic in Chiredzi town, more than 20 km away. No recorded interventions exist in Chipimbi, and data on infection prevalence in the community is severely lacking. The community relies primarily on open water sources such as the Chipimbi River and Magumire Dam for daily activities related to agriculture, fishing, and personal use. Both *Biomphalaria* and *Bulinus* spp. were recently reported in the Chipimbi River [20], with the former found to be infected with *Schistosoma mansoni* at an infection prevalence of 11.1% (2/18).

Despite the proximity of HVE and Chipimbi and their shared climatic and cultural influences, the stark differences in access to water, sanitation, and healthcare facilities provide a compelling comparative setting for this study.

### Ethics statement

Formal consent to carry out this study in the designated area was granted in writing by the highest regional authority, the office of the District Administrator, and ethical clearance was secured from the Social and Societal Ethics Committee at the University of Leuven (ref. no. G-2022-5508-R2(MIN)).

In adherence to customary practices within Zimbabwean rural communities, explicit permission was also sought from the village heads, complementing the approval granted by the District Administrator. Specifically, within Chipimbi, a region further segmented into distinct villages, the consent process extended to the Crow Heads with whom a meeting was organized regarding the authorization, ensuring comprehensive community involvement and adherence to local protocols.

Participants were provided with comprehensive oral and written information detailing the nature and purpose of the study. Prior to their inclusion, individuals expressed their understanding and agreement by signing a written informed consent document. To safeguard confidentiality and anonymity, a unique code was assigned to each participant, dissociating personal identifiers from the study data.

### Quantitative approach through questionnaires

Pre-structured questionnaires were employed to assess the KAP levels of schistosomiasis. Initially developed in English (https://ee.kobotoolbox.org/x/GxcBYKog), the questions were later translated into Shona (https://ee.kobotoolbox.org/x/g4hcTxJd). KoboToolbox’s online platform facilitated immediate digitization of responses, accessible to the research team via the KoboCollect application. Other advantages included features like skip-logic and mandatory questions. The questionnaire covered demographic, socio-economic, and environmental data, alongside practices related to schistosomiasis, including personal hygiene, water contact, and treatment history. Additionally, questions on schistosomiasis knowledge encompassed etiology, transmission, clinical manifestations, prevention, and control, incorporating closed-ended and open-ended formats to capture quantitative and qualitative data. Specifically, knowledge-related questions were open-ended, without multiple-choice answers which might have given hints to the respondent thereby preventing potential biases. However, practice-related questions presented multiple-choice options for assessing the frequency of the participant’s choice of these activities or actions.

The questionnaire comprised 72 questions, covering 15 on demography, 12 on knowledge, 15 on attitudes, 13 on practices, and 17 on infection, prevention, and health-seeking behavior (Fig 2). Demographic questions aimed at general information, knowledge questions assessed respondents’ understanding of schistosomiasis, attitude questions gauged participants’ seriousness and willingness to change behavior, and the practices section focused on actions predisposing participants to infection. Utilizing skip-logic resulted in varied question numbers per respondent based on given answers. Instead of schistosomiasis, the questionnaire used ‘*bilharzia*’ to align with local terminology.

**Fig. 2.**
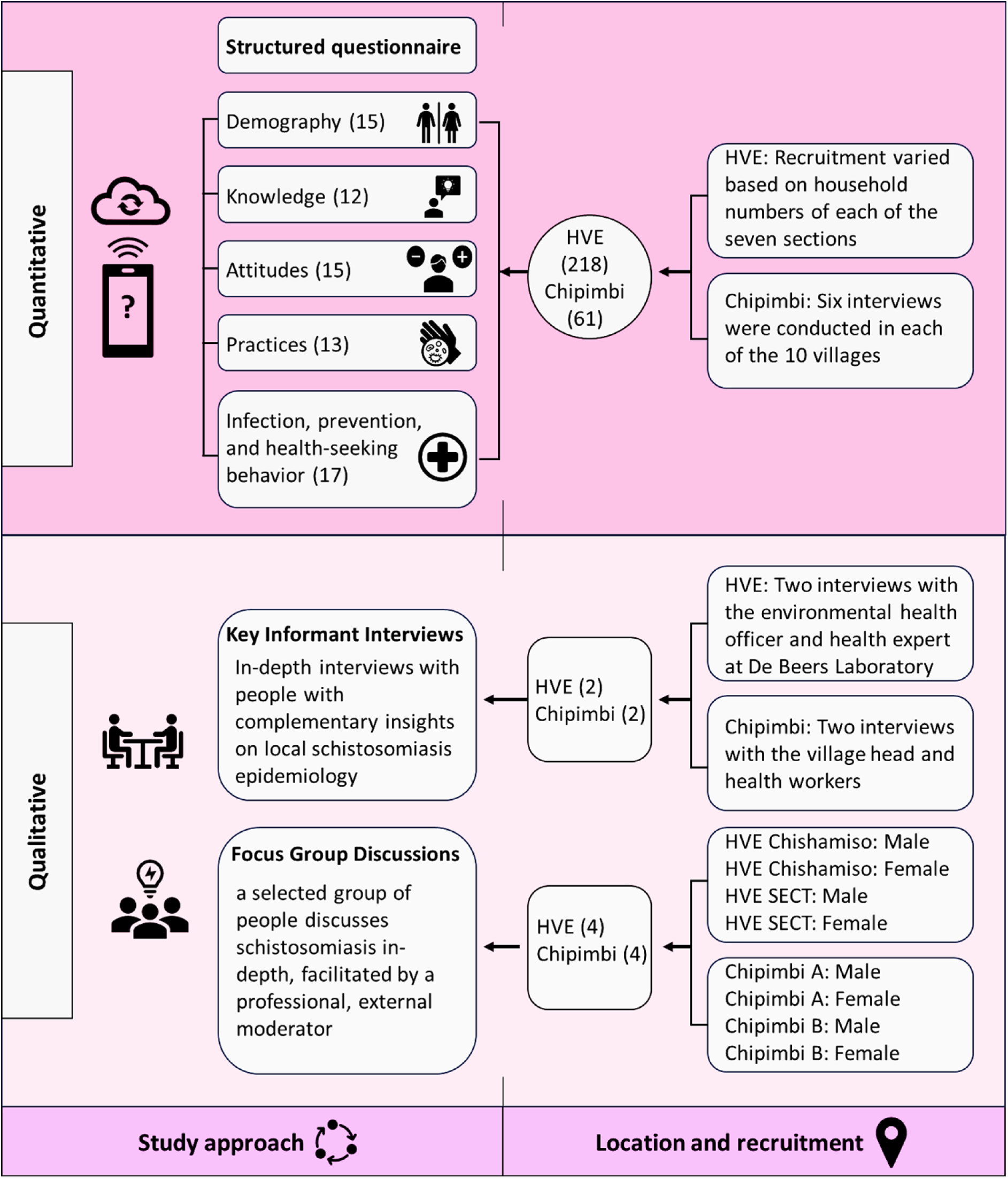
Schematics of the study methodology, location, and recruitment of respondents and participants.

The sample size for our study was determined using Kish’s formula (n = Z² PQ/E²). With a Z-score of 1.96 (95% confidence interval) and a schistosomiasis prevalence of 22.7% in Zimbabwe [8]. The calculated sample size was 269, distributed between HVE and Chipimbi in a 3:1 ratio. Q is the proportion not infected, and E is the standard error set at 0.05. Ultimately, 279 questionnaires were administered – 218 in HVE and 61 in Chipimbi. To ensure representation, at least six interviews were conducted in each of the 10 villages in Chipimbi. In HVE, considering varying household numbers across sections, a similar per-section calculation was employed (Additional Table 1).

A blend of stratified and systematic sampling ensured geographic coverage and demographic representation. Starting randomly, interviews proceeded at fixed intervals. Participants, aged 18 or older, were limited to one per household to avoid correlated responses. Interview intervals were determined by dividing the (estimated) number of households per village or section by the required questionnaires.

The research team consisted of the principal investigator (female), field supervisors (two males, one female), and community research assistants (CRAs) (two males, eight females). The CRAs were recruited from their respective HVE sections and all had prior experience in conducting similar surveys. They received additional training on the technical aspects of the questionnaire and the practical use of KoboToolbox. To ensure strict adherence to the sampling design, the geo-location of each sampled household was captured in real-time using KoboToolbox. Data from KoboToolbox underwent cleaning in Microsoft 365 Excel before export to IBM SPSS Statistics for analysis.

### Qualitative approach through focus group discussions

Eight focus-group discussions (FGDs) enriched quantitative data by exploring motivations. FGDs, involving eight to 12 participants per session, following Khan et al [21], lasted one to one and half hours. Sessions were single-sex (male or female) representing a wide age range and the various villages (Fig 2). Gender-based FGDs aimed to capture diverse KAPs on schistosomiasis due to potential differences in gender roles. Participants, fluent in Shona, were selected by community leaders and health workers to ensure full representation of the different villages.

### Qualitative approach through key informant interviews

Key informants, chosen for their central community roles or schistosomiasis expertise, underwent semi-structured interviews (Additional Text 1). Four face-to-face KIIs were conducted, featuring the Chipimbi village head, two Chipimbi healthcare workers, a health researcher from the Chiredzi district, and the senior Environmental Health Officer of HVE. The aim was to understand information on past treatment campaigns, available WASH and previous WASH campaigns, presence and capacity of hospitals, and to gauge community knowledge.

### Quantitative data analysis

The study considered six independent variables: age, education level, monthly income, marital status, gender, and residence. For statistical analysis, these variables were assigned numerical values. Ordinal variables such as age (categorized as 18-27, 28-37, 38-47, 48-57, 58-67, and 68+ years), education level (none, primary, lower secondary, higher secondary, tertiary), and monthly income (less than 50 USD, 51-100 USD, 101-150 USD, 151-200 USD, above 200 USD) were recoded, with the lowest category assigned a value of 1, the second lowest as 2, and so on. Residence, marital status, and gender were treated as binary variables.

The knowledge, attitude, and practice (KAP) sections were scored as follows:

i. Knowledge: Each correct response in the knowledge section received 1 point, with a maximum possible score of 40. The knowledge score was computed by summing the correct answers. Scores ranged from 0 to 40, with knowledge levels categorized as low (0-13 points, coded as 0), moderate (14-26 points, coded as 1), and high (27-40 points, coded as 2).
ii. Attitude: Likert scale responses were used to measure attitude, with scores ranging from 1 to 5. Thirteen statements were averaged to compute the overall attitude score, categorized as follows: negative attitude (1.00-1.99), somewhat negative attitude (2.00-2.99), somewhat positive attitude (3.00-3.99), and positive attitude (4.00-5.00). The internal consistency of the Likert scale was assessed using Cronbach’s alpha (α) coefficient, with the following interpretation: ≥ 0.9 = excellent, ≥ 0.8 = good, ≥ 0.7 = acceptable, ≥ 0.6 = questionable, ≥ 0.5 = poor, and ≤ 0.5 = unacceptable [22].
iii. Practice: Responses indicating safe practices related to water use and sanitation were awarded 1 point, with a maximum possible score of 14. Practice levels were categorized as bad (1-4 points), moderate (5-9 points), and good (10-14 points).

Descriptive statistics, including frequency values and percentages, were generated for the KAP variables and the sociodemographic profile of respondents. Contingency tables were created, and Cramer’s V (φ) and Gamma (γ) coefficients were calculated to determine the associations between sociodemographic characteristics and KAP variables, following the methodology of Wambui et al [22]. A p-value of 0.05 was used to establish statistical significance. The strength of associations was interpreted using the rule of thumb from Wambui et al [22]: 0.00-0.10 (negligible), 0.10-0.30 (small), 0.30-0.50 (medium), and 0.50 or more (large). All statistical analyses were performed using IBM SPSS Statistics version 28.0.1.1 (14).

### Qualitative data analysis

Data from FGDs and KIIs were translated and transcribed to English using the oTranscribe online platform (https://otranscribe.com/). The framework method described by Gale et al [23] was used to analyze the qualitative data. Initially, text was transcribed, then carefully read and coded, with codes representing information types. Deductive coding, based on clear research objectives and expected discussion outcomes, incorporated pre-defined codes derived from questionnaire questions and answers. Additional codes were introduced as needed. Codes were categorized into main and subcategories, allowing for easier comparisons between the different texts. NVivo Release 1.7.1 (1534) was used to manage and process the qualitative analyses. To ensure robustness, only statements and themes mentioned in at least two different groups or by multiple FGD members were included in the results, minimizing reliance on individual views unsupported by others.

## Results

### Sociodemographic traits of respondents

Hippo Valley Estates housed the majority of respondents (78%), with over half being female (56%). Half were aged 37 or below (50%), while only 1% were older than 68. Around two-thirds were married (59%), either monogamously or polygamously (6%). One-third belonged to households of three to four people (33%) or five to six people (33%). Nearly half had lower secondary education (49%), and almost all were literate (95%). Over 60% earned less than $50 per month. Crop growing was the predominant occupation (23%), while about 18% were unemployed, 14% were employed in the informal sector. Various religions were represented, with Christian Pentecostal being the most common (29%) followed by the African apostolics (25%), Catholics (11%) and the African indigenous practitioners (10%). The majority identified as Shona (73%) followed by Shangani (15%). Additional Table 2 has more details on sociodemographic traits of respondents.

### Knowledge with regards to schistosomiasis

A large proportion of respondents in both HVE (88%) and Chipimbi (85%) were aware of schistosomiasis. Almost all recognized it as a disease (98%), and most identified snails as the transmission organism (HVE: 77%, Chipimbi: 57%). In HVE, many correctly identified transmission routes, with swimming and bathing (89%) being most mentioned, followed by irrigation work (43%) and fishing (35%). Drinking untreated water was also linked to transmission (47%). Chipimbi participants most frequently associated transmission with swimming and bathing (75%) and drinking untreated water (71%).

Several misconceptions about schistosomiasis were identified. In Chipimbi, 31% of respondents mistakenly believed that schistosomiasis could be avoided by washing fruits and vegetables, while 33% thought it could be prevented by avoiding walking barefoot. These misconceptions were less prevalent in HVE, where only 14% and 11% of respondents, respectively, held these views. Other incorrect beliefs included the association of schistosomiasis transmission with playing in the soil (HVE: 5%, Chipimbi: 2%) and sexual contact (HVE: 2%, Chipimbi: 6%).

These misconceptions were also highlighted in the focus group discussions (FGDs). For example:

> *“Also teach them [children] that when they go to the toilet, they should not urinate on the floor. And that they should always wear shoes, so they don’t get bilharzia”* – **female, HVE section 24.**

> *“The way bilharzia is painful, as mentioned by the previous speaker, might be like STI because for us women when it’s bilharzia, this part here burns [pointing to the loins]. You will finally release urine after a lot of pain. Siki [local name for STIs] also shows the same burning sensation and signs. So, when you try to explain it as bilharzia to someone else, they will say you have siki and will not think of bilharzia”* – **female, Chipimbi 1**.

Further misconceptions came up in the FGDs where participants associated schistosomiasis with eating too much salt:

> *“We also hear in this community that if children eat a lot of salt it causes them bilharzia.”* – **female, HVE section 24.**

> *“We would hear or mothers and grandmothers say: ‘do not play in the sun, because if you play in the heat you will get bilharzia. Also do not eat a lot of salt, otherwise you get bilharzia’”*. – **female, Chipimbi 2**.

Commonly associated symptoms were blood in urine (HVE: 83%, Chipimbi: 98%), painful urination (HVE: 63%, Chipimbi: 87%), and blood in stool (HVE: 47%, Chipimbi: 50%). Complications like bladder and kidney damage (HVE: 65%, Chipimbi: 60%), infertility (HVE: 47%, Chipimbi: 54%), and increased prostate cancer risk (around 40% for both communities) were also recognized (Additional Table 3).

Over half of the respondents recognized water source treatment as a preventive measure for schistosomiasis (HVE: 61%, Chipimbi: 75%), followed by avoiding open defecation and urination (HVE: 58%, Chipimbi: 64%) and treatment for all infected individuals (HVE: 37%, Chipimbi: 65%). Almost all participants acknowledged the role of urinating and defecating by water in spreading schistosomiasis (HVE: 93%, Chipimbi: 88%). Nearly all respondents were aware that drugs from health centers, pharmacies, or over-the-counter can help cure schistosomiasis (HVE: 96%, Chipimbi: 98%) (Additional Table 3).

Ultimately, many respondents could be classified as having poor to moderate knowledge with regards to schistosomiasis in both regions. Gamma (γ) and Cramer’s V (φ) were employed to assess the association between schistosomiasis knowledge and sociodemographic characteristics of the respondents (Table 1). A moderate, positive association was identified between monthly income and the knowledge score (γ = 0.273, p = 0.005), suggesting that respondents with higher incomes generally possess greater knowledge of schistosomiasis. The community of residence (φ = 0.212, p = 0.002) and gender (γ = 0.183, p = 0.009) also exhibited a small but significant association with knowledge of schistosomiasis. The direction of association could not be established due to the nature of the variable community or the variable gender and the statistical test.

**Table 1.** Knowledge, attitudes and practices score against age, education, monthly income, community, gender, and marital status.

| General knowledge score | Age |  | Education |  | Monthly Income |  |
| --- | --- | --- | --- | --- | --- | --- |
| | $\gamma$ | P value | $\gamma$ | P value | $\gamma$ | P value |
|  | .026 | .754 | .162 | .076 | **.273** | **.005** |
|  | Community |  | Gender |  | Marital status |  |
| | $\phi$ | P value | $\phi$ | P value | $\phi$ | P value |
|  | **.212** | **.002** | **.183** | **.009** | .054 | .666 |
| General attitude score | Age |  | Education |  | Monthly Income |  |
|  | Y | P value | Y | P value | Y | P value |
|  | **.145** | **.012** | .024 | .700 | **.169** | **.015** |
|  | Community |  | Gender |  | Marital status |  |
| | $\phi$ | P value | $\phi$ | P value | $\phi$ | P value |
|  | **.611** | **<.001** | .277 | .436 | .316 | .144 |
| General practice score | Age |  | Education |  | Monthly Income |  |
|  | Y | P value | Y | P value | Y | P value |
|  | .088 | .127 | .116 | .078 | **.238** | **.001** |
|  | Community |  | Gender |  | Marital status |  |
| | $\Phi$ | P value | $\phi$ | P value | $\phi$ | P value |
|  | **.385** | **<.001** | .256 | .051 | .219 | .206 |

### Local attitude towards schistosomiasis

A general agreement existed between respondents in HVE and Chipimbi that indeed schistosomiasis is a serious disease (Fig. 3). Respondents reflected it is necessary to take preventative measures against the disease (HVE: 98%, Chipimbi: 100%), it is important to know if they have schistosomiasis (HVE: 97%, Chipimbi: 97%), and that they will take appropriate action if they discover they are infected (HVE: 97%, Chipimbi: 98%). The majority (highly) agreed that the disease could be cured (HVE: 94%, Chipimbi: 98%) and realized the importance of avoiding water contact (HVE: 80%, Chipimbi: 86%). Almost all respondents (highly) agreed with the statement that urinating and defecating in the toilet is important for their health (HVE: 98%, Chipimbi: 98%). The statement that taking medication for schistosomiasis is important for their health was also (highly) agreed upon by many respondents (HVE: 95%, Chipimbi: 100%). The respondents (highly) agreed that they should go to the hospital if they find they have schistosomiasis (HVE: 99.5%). In HVE, a few people (highly) agreed they should go to a traditional health practitioner (13%) or religious leader (14%) when they find they are infected with the disease, while for Chipimbi it was 10% and 16% respectively. Most of the respondents (highly) agreed that it is important to be informed about schistosomiasis (HVE: 95%, Chipimbi: 100%) and that they would wish to get more information on it (HVE: 97%, Chipimbi: 100%).

**Fig. 3.**
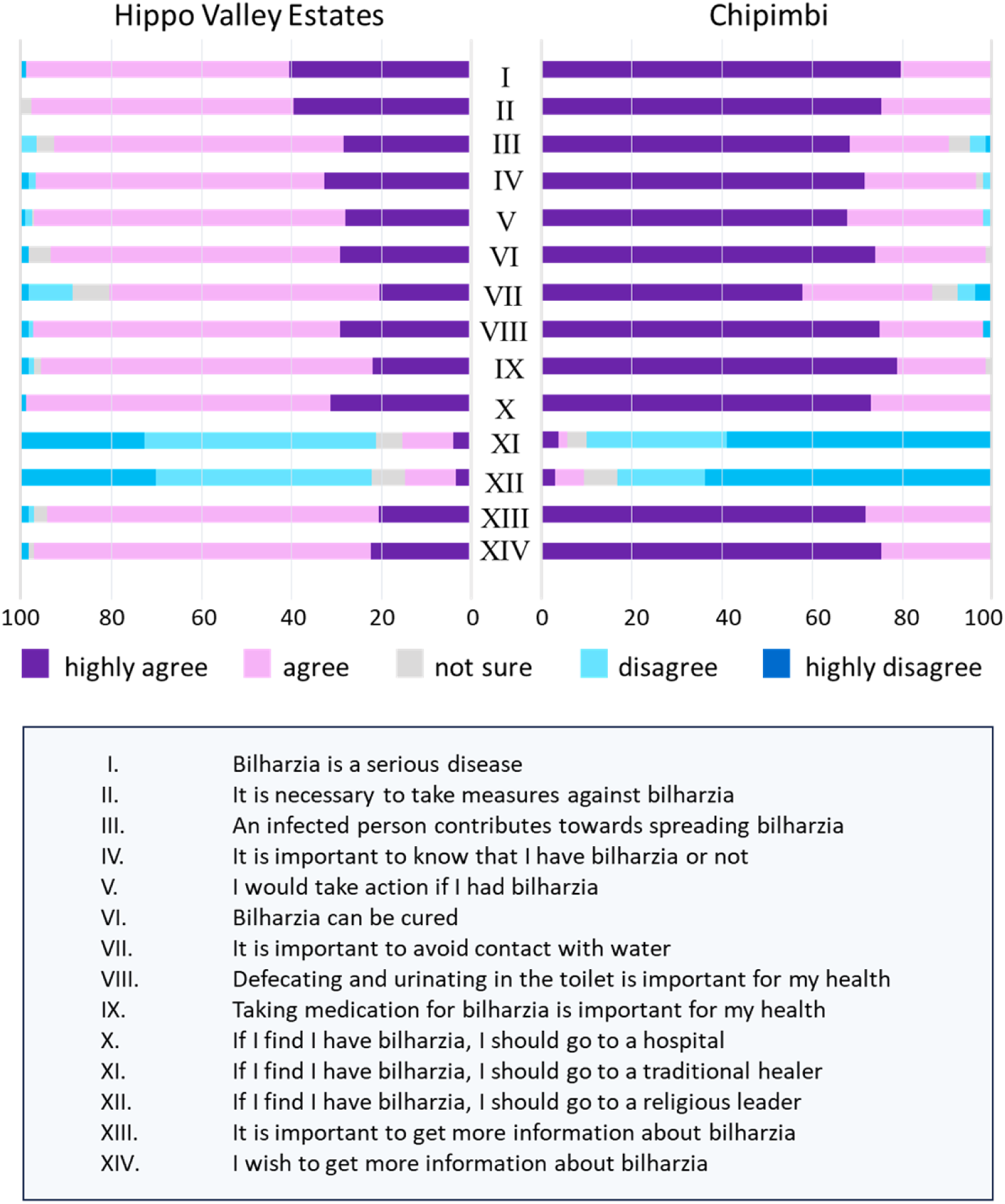
Attitudes of Hippo Valley Estates and Chipimbi community members toward schistosomiasis.

At least one participant in each FGD – aside from the female group at HVE Chishamiso – believes that schistosomiasis is a serious illness. Participants linked the seriousness of the disease to the possible long-term consequence of becoming infertile and adverse effects on child development:

> *“It [bilharzia] might be a very serious disease because you might fail to get children eventually, in the future”* – **male, HVE section 24.**

> *“It is a very important disease because in our lives when we are growing up, we are expecting to bear children. So, when it causes me not to have children, it becomes a very important disease to me.”* **female, Chipimbi 2**.

> *“When we have children, we expect them to grow up and become nurses, doctors, teachers, but when they get infected, there is nothing good that will come out of them. This means that as a nation or family, we would have lost the potential of this child. They could have become nurses, doctors, teachers, or even something better like president, it would have helped the nation to become better in the future, but all is gone.”* – **female, Chipimbi 2**.

However, not all participants shared this sentiment; others pointed out that schistosomiasis is not a disease that people in their village or section take seriously.

> *“Of course, it’s a disease that occurs but it is not very scary because even when you get it you can be cured. Right?”* – **male, Chipimbi 1**.

> *“When we were growing up, this disease was regarded as very important. Our mothers would tell us that ‘you don’t play in the water. The stagnant water – never play in that water!’. But now with other new, more sophisticated diseases, this disease has been overshadowed by many other diseases. And today no one even talks about it.”* – **female HVE section 24**.

The gamma coefficient (γ) and Cramer’s V (φ) were utilized to assess the association between sociodemographic characteristics and attitudes (Table 1). Both age (γ = 0.145, p = 0.012) and monthly income (γ = 0.169, p = 0.015) demonstrated a small positive association with respondents’ attitudes toward schistosomiasis. This suggests that older respondents and those with higher monthly incomes are more likely to harbor a more positive attitude toward schistosomiasis. Additionally, a strong and significant association was observed between the community of residence of the respondent (φ = 0.611, p = <0.001) and their attitudes.

### Health-seeking practices and local access to healthcare

Some respondents highlighted that someone in their household suffered from schistosomiasis (HVE: 29.8%, Chipimbi: 39.3%) and most patients consulted a health facility (HVE: 87.7%, Chipimbi: 95.8%). While a significant portion of HVE (72.5%) took proactive measures to prevent bilharzia, a somewhat lower percentage in Chipimbi agreed to do so (55.7%), with some citing a lack of knowledge on preventive measures (Table 1). The most frequently mentioned prevention method was avoiding open defecation (HVE: 75.3%, Chipimbi: 70.6%), followed by medication (HVE: 54.4%, Chipimbi: 47.1%). Avoiding contact with water was mentioned less frequently (HVE: 12%, Chipimbi: 17.6%). A substantial percentage of respondents in HVE (77.5%) had received praziquantel before, while nearly half in Chipimbi had done so (54.1%). Reasons for not receiving praziquantel included the absence of a campaign or lack of awareness about the campaign. Most respondents were open and receptive to a vaccine against schistosomiasis (HVE: 83.9%, Chipimbi: 93.4%). Additional Table 4 gives a full description of health-seeking practices towards schistosomiasis.

FGDs held in HVE and Chipimbi showed that most respondents would go to a hospital if they noticed they or their children were infected. Several participants of all discussions expressed their faith in professional healthcare even above the local traditional treatment options:

> *“Soon after seeing that you have the disease, waste no more time but run to the hospital and describe it as it is and they will examine you”* – **male, Chipimbi 2**.

> *“People now quickly seek medical health at the hospital”* – **KII Village Head Chipimbi 2**.

> *“At the traditional healer, I don’t think it [bilharzia] will completely go away. Yes it might reduce a bit like the previous speaker said, but to say it is completely cured, that is not going to happen. Only the hospital can cure it”* – **male, HVE section 24.**

Several factors were mentioned to be confounding to the access to primary health care in both communities. The first was selective access to health care. Tongaat Hulett supplied a hospital and clinics, according to the environmental health officer in the KII, but FGDs show that these facilities were exclusively intended for HVE employees and their families, excluding contract laborers and others not affiliated with Tongaat Hulett.

> *“As long as you do not have a company number, you will never receive treatment in this place.”* **-male, HVE Chishamiso.**

Second, some health facilities were reported to be under-equipped with both medical and human resources. When residents of Chipimbi come to them with signs and symptoms, the VHWs of Chipimbi 1 mentioned in the KII that they cannot assist them beyond referring them to the hospital. Nevertheless, it is said that the hospitals are devoid of medication.

> *“We refer the patient to the hospital. And when they go to the hospital, the nurses there will examine them and try to find out what exactly bothers them.”* – **KII VHW Chipimbi 1**.

> *“You might not find it [the medication] there, but most importantly you might fail to raise the transport money to get there”* – **male, Chipimbi 2**.

> *“Sometimes you cannot find the medicine, even if you go to the hospital. They’ll tell you that the medicine is not there. And now you are left to look for other money to get the bus to go to the general hospital of Chiredzi. And when you get to the general hospital, they also tell you that they don’t have the medicine. Now I must go to the pharmacist. Because I don’t have money, I am forced to come back home and continue to live with the disease”.* – **female, Chipimbi 2**.

Third, the difficulty of getting to the clinic or hospital was brought up in every Chipimbi FGD. The distance to the nearest hospitals, Chizvirizvi Rural and Chipiwa Hospitals ranges from 10 to 40 km, depending on the place of residence within the villages. As a result, some participants in Chipimbi opt for self-treatment or traditional healing and only go to the hospital when such methods do not work:

> *“They have assigned Chipiwa as our hospital, but it is still far away, and you would need a bus to get there.”* – **female, Chipimbi 1**.

> *“When I know that I am in trouble, before I think about the hospital I will first think, ‘Can a certain herb help me?’ Then I dig it up and consume it. Maybe it might cure me but if it persists then I go to the hospital”* – **male, Chipimbi 1**.

Fourthly, several females in HVE Chishamiso raised the issue of how they were afraid that their medical information would be shared with the community, and this led to some of them avoiding seeking professional healthcare. One FGD participant would resort to a traditional healer:

> *“I would start with seeing the traditional healers first and get the necessary herbs and go here [the local clinic] only when it doesn’t work with the traditional healer. Because the fear is with the health workers here in this community, when you tell them, your medical health issues will be spread throughout the entire community. But the traditional healers will keep my secrets.”* – **female, HVE Chishamiso**.

### Risky practices and water access

The majority of HVE respondents identified piped water as their primary household water source (98%), with only a small percentage (6%) mentioning natural sources like rivers, streams, lakes, ponds, or irrigation canals. Water-related activities reported included bathing (87%), laundry (81%), and water fetching (75%), with 51% visiting the water source more than three times daily. Despite a general presence of taps in most homes, the environmental health officer and participants in all HVE FGDs noted inconsistent water availability.

> *“We make sure that it [tap water] is adequate, it is treated, it is always available, and it is free for the users.”* – **KII, environmental health officer in HVE**.

> *“When it [tap water] runs out when it finishes, we would go and fetch in the canal.”* – **female HVE Section 24.**

> *“When there are power cuts, when there is no electricity, the water doesn’t come out from the taps, and you have to go to the canals to take a bath.”* – **male HVE Chishamiso**.

In Chipimbi, the primary sources of household water, as reported by most respondents, were boreholes (97%) and natural sources like rivers, streams, lakes, ponds, or irrigation canals (46%). The village head of Chipimbi 2, in a KII, also emphasized the utilization of boreholes, rivers, and dams. Village health workers mentioned in the KII that some boreholes were funded and drilled by the Chinese through China Aid. Respondents highlighted various water-related activities, including bathing and laundry (100%), water fetching (95%), agriculture (84%), taking animals for drinking (54%), and fishing (54%). The frequency of visits to water sources was reported with most respondents indicating twice (30%) or thrice a day (26%). FGDs further elaborated on the use of water from different water sources.

> *“At the borehole, they [women] get one bucket to drink, the rest [doing laundry, etc.] is from the dam”* – **male, Chipimbi 1**:

> *“We have a borehole and a tap, but we also have a river. That river is very important to us. That is where we get water to wash and we wash while we are standing in that water, where they say the bilharzia worms are found.”* – **female, Chipimbi 2**.

Risky practices by children have been discussed in three KII:

> *“They [children aged 11-15] just come and swim [in the dam and river], but when they swim, because children normally don’t know, they’ll just be thinking to themselves ‘Oh I am just having fun, I am just taking a swim’, but without knowing that in that swimming there is a chance of catching some germs that are already there in the water.”* – **KII, VHW Chipimbi 1**.

> *“You will find they [school-aged children] would want to play in shallow canals.”* – **KII, environmental health officer in HVE**.

In the KII with the environmental health officer of HVE, it was established that the Tongaat Hullet organization in charge of HVE is aware employees are at risk of schistosomiasis infection, due to the intensive use of water. Engineering programs are in place to alleviate this:

> *“Our way of doing business creates risk for bilharzia transmission…. we are trying, by all means, to make sure that our people are safe…. through a fortnightly program where all waterbodies are inspected and treated to control the vectors…. a majority of the canals are cement lined to allow water to flow and discourage survival of vector snails.”* – **KII, environmental health officer in HVE**.

### Practices related to access to and use of toilets

In HVE, nearly all respondents, except two, reported that their households have a toilet, and the majority use a flush system (92%). About half of the respondents admitted to occasionally defecating or urinating outside the toilet (51%), with the majority choosing the bush as the preferred location (81%), while a small percentage did so near water bodies (9%). Less than half of the respondents mentioned rare instances of such behavior (46%), while one in five reported doing so quite often (20%). The primary reasons cited were the urgency of nature’s call (53%), lack of access to toilets (26%), and insufficient water to flush the toilet (16%).

Based on the insights gathered from the FGDs in HVE, most people generally have access to either private or shared communal toilets. Participants have, however, reported using the canal or bush instead of the toilets due to several issues such as the availability of water to flush down the toilets.

> *“Sometimes you might feel very lazy to go to the canal to get water to use for the toilet and simply just go to the bushes directly to defecate there.”* – **female, HVE section 24.**

> *“If people get there [the toilet] and they find out that the toilet is dirty they’ll go to use the canal. Or they don’t use the toilet hole, they will do it on the floor instead.”* – **female, HVE Chishamiso**.

In Chipimbi, a significant portion of respondents utilized pit latrines (48%), followed by improved pit latrines (26%), while 26% indicated that their households had no toilet facilities. A considerable number of respondents admitted to defecating or urinating outside the toilet (62%), citing reasons such as nature calls (66%) and lack of access to toilets (50%). The majority of those going outside the toilet did so in the bush (74%). Among those mentioning such behavior, approximately one-third did so rarely (32%), while 29% reported doing so quite often. Discussions in all FGDs and KIIs in Chipimbi highlighted the scarcity of available toilets and this seems to have been understated in the questionnaires.

> *“People without toilets are more than the ones that have toilets.”* – **KII, Village Head Chipimbi 2**.

> *“In village 1 there are only nine toilets, out of 84 households. In village 2 there are 50 households and only 12 toilets.”* – **KII VHW Chipimbi 1**.

> *“Most of us go to the bush because we do not have toilets.”* – **female, Chipimbi 1**.

> *“Everyone wants a toilet but lacks money.”* – **female, Chipimbi 1**.

In the FGDs, open defecation was reported to involve men, women, and children, although it appears to be more prevalent among children.

> *“Everyone is involved—children, men, and women. Someone might be washing themselves in the canal and think, ‘Do I really have to get out of the canal to defecate and then come back to wash again?’ So, they just let it flow downstream.”* – **Female, HVE Chishamiso**.

> *“Mostly children. But as mentioned, some men are also involved in these practices. Even some adults, who can be quite difficult to correct, engage in such behavior. They can be very mischievous, so they might end up doing that.”* – **Male, HVE Section 24.**

### Association between sociodemographic characteristics and risky practice scores

Overall, a small, positive association is found between the monthly income of a respondent and their score on practices related to behavior putting themselves at risk of schistosomiasis (γ = 0.238, p=0.001) (Table 1). The positive association implies that respondents with a higher monthly income are more likely to have better practices that put them less at risk of schistosomiasis. A moderate, significant association was also found between the community of residence of a respondent and their practice score (φ =0.385, p = <0.001), implying that the community of residence influences how respondents engage in practices that are minimizing schistosomiasis transmission.

## Discussion

We employed a mixed-methods approach to explore the knowledge, attitudes, and practices surrounding schistosomiasis in two distinct communities within the Chiredzi district of Zimbabwe, each differing in their WASH (Water, Sanitation, and Hygiene) facilities and interventions. The study targeted respondents aged 18 and above, encompassing a diverse range of ages, occupations, genders, and residential locations. By combining quantitative and qualitative techniques – including focus group discussions (FGDs), key informant interviews, and questionnaires – we gathered comprehensive and complementary data. This approach provided a nuanced understanding of the disparities in facilities and interventions between Hippo Valley Estate (HVE) and Chipimbi, as well as the challenges these communities face. Here, we discuss how these differences could impact infection risk and the effectiveness of current and future control initiatives.

### Widespread awareness, but gaps in schistosomiasis knowledge persist

Our study revealed high levels of awareness about schistosomiasis among participants, with 88% from Hippo Valley Estate (HVE) and 85% from Chipimbi having heard of the disease, and 98% recognizing it as a disease. These findings align with similar studies from sub-Saharan Africa, where awareness levels were reported as 91% in Mozambique [24], 91% in South Africa [25], 79% in Gabon [26], 91% in the DRC [22], and 99% in Uganda [27]. Within Zimbabwe, high awareness levels were also reported in Gwanda (81%) [25] and Shamva (94%) [15].

A moderate positive association was found between monthly income and knowledge of schistosomiasis (γ = 0.273, p = 0.005), suggesting that higher-income respondents generally had greater knowledge of the disease. This may be attributed to better access to educational resources among wealthier individuals. Additionally, community of residence (φ = 0.212, p = 0.002) showed a small but significant association with schistosomiasis knowledge, likely reflecting the more extensive health education and intervention efforts historically present in HVE, as reviewed by Chimbari [28].

Most participants first learned about bilharzia in school (HVE: 65%, Chipimbi: 44%), followed by healthcare workers (HVE: 15%, Chipimbi: 23%). These sources were also highlighted in focus group discussions, emphasizing the critical role of schools and healthcare workers in raising awareness. The government of Zimbabwe has recently launched the School Health Policy in 2018, a framework through which the central role of schools in teaching health related issues will increase [29]. As also outlined in the Zimbabwe National Master Plan for the Elimination of Neglected Tropical Diseases for the years 2023 to 2027, behaviour change interventions will need to target schools given that children are more susceptible to NTDs and are effective change agents in households and local communities [30]. Further, a majority, particularly in Chipimbi (64%), recognize health education as a measure for schistosomiasis control and prevention. Therefore, it would be beneficial to include prudent curriculum on schistosomiasis especially also because mass drug administration campaigns are already included in this policy.

Knowledge about the transmission of schistosomiasis and the role of snails was also high among participants (HVE: 88%, Chipimbi: 67%), which is significantly greater than in a similar study in Kenya, where only 3% made this connection [31]. However, few participants recognized the importance of snail control in disease prevention. Many respondents understood the role of contact with contaminated water in transmission (HVE: 89%, Chipimbi: 75%), consistent with findings from Uganda (81%) [27] and higher than in Shamva, Zimbabwe (44%) [15].

Despite this, some myths and misconceptions persist. In Chipimbi, 31% of respondents believed schistosomiasis could be avoided by washing fruits and vegetables, and 33% by avoiding walking barefoot, views held by very few in HVE (14% and 11% respectively). Focus group discussions further revealed misunderstandings, such as the belief that schistosomiasis can be contracted through direct contact with the urine of an infected person, particularly by entering a toilet barefoot. This mirrors findings from Gwanda, Zimbabwe [25]. While it is true that entering toilets barefoot does not transmit schistosomiasis, it does increase the risk of infection with soil-transmitted helminths, which are also prevalent in the region [8,32]. The historical overlap of treatment campaigns for schistosomiasis and soil-transmitted helminths in Zimbabwe may have contributed to confusion regarding the transmission of these parasites.

Moreover, participants in four out of eight focus groups associated eating too much salt with bilharzia. This misconception is not unique to Zimbabwe; a study in Kano State, Nigeria, found that some people believe eating salty or sour foods causes schistosomiasis [33]. We speculate that this myth may stem from the logic that excessive salt intake leads to water retention, resulting in low volumes of concentrated, darker urine – resembling the blood in urine that is a common symptom of urinary schistosomiasis. Although this is speculative, it offers a plausible explanation for the persistence of this myth in geographically and ethnically diverse regions.

In 60% of reviewed studies, schistosomiasis was mistakenly believed to be sexually transmitted [4], with 27% in Mozambique, 26% Nigeria [34], and 27% in the DRC [22]. However, in our study, only 2% of HVE and 6% of Chipimbi participants held this belief, consistent with findings from South Africa and Zimbabwe [25]. Nonetheless, some women in both communities noted that the presence of blood in urine – a common symptom of schistosomiasis – could be confused with sexually transmitted infections (STIs), potentially leading to reluctance in reporting symptoms. This was not discussed in the male groups. This gender difference might be because of uneven health and social consequences related to STDs for women compared to men [35].

The most commonly reported symptoms of schistosomiasis were blood in urine (HVE: 83%, Chipimbi: 98%) and painful urination (HVE: 63%, Chipimbi: 87%), consistent with studies from Zimbabwe (71%) [15], South Africa (65%) [36], and Gabon (91%) [26]. These symptoms are indicative of urinary schistosomiasis [3]. The long-term consequences of untreated schistosomiasis, such as kidney and bladder damage, and infertility, were frequently mentioned in both questionnaires and focus group discussions. The emphasis on urinary symptoms, despite the presence of both urinary and intestinal schistosomiasis in the study area, may be due to the more distinctive nature of blood in urine compared to gastrointestinal symptoms, which are common to other infections. This disparity may loosely indicate a situation of underreported cases of intestinal schistosomiasis, underscoring the imperative to incorporate this aspect into forthcoming health education programs.

### Attitudes regarding schistosomiasis

Attitudes toward schistosomiasis play a critical role in determining how far local communities will go to protect their health and the potential success of disease control initiatives. In this study, nearly all respondents regarded schistosomiasis as a serious disease (HVE: 99%, Chipimbi: 100%). This aligns with recent findings from Uganda, where 97% of participants held similarly serious views about the disease [27]. Generally, respondents exhibited positive attitudes towards schistosomiasis and its treatment, with older participants showing stronger attitudes (γ = 0.145, p = 0.012), consistent with observations from Brazil [37]. Interestingly, a strong association was found between respondents’ community of residence and their attitudes (φ = 0.611, p < 0.001), with Chipimbi displaying more positive attitudes than HVE, despite HVE’s history of more extensive interventions.

Focus group discussions (FGDs) in this study revealed that the perceived seriousness of schistosomiasis was often linked to its long-term consequence of infertility. In many African societies, fertility is highly valued, both at the individual and community levels, fulfilling cultural and religious expectations [38–40]. For instance, Shona women in Zimbabwe often define themselves by their marital status and motherhood [41], while in Eswatini, a woman’s fertility enhances her status within her marital family [42].

However, not all participants believed that people in the two communities were taking schistosomiasis seriously. This sentiment mirrors findings from the Democratic Republic of Congo (DRC), where the disease is often not regarded as serious, likely due to a lack of knowledge about its link to fertility [22]. As highlighted by FGD participants, good knowledge fosters positive attitudes, yet the perceived seriousness of schistosomiasis may be diminished by the emergence of more threatening diseases like HIV/AIDS, malaria, and COVID-19. Diseases like schistosomiasis, which often have subtle or unnoticed symptoms (e.g., bladder fibrosis and anemia), struggle to be taken seriously in communities facing multiple challenges – an issue likely exacerbated in the DRC, where average income is lower than in Zimbabwe. This context may contribute to schistosomiasis being an ‘accepted tropical disease’ that is overlooked.

Other reasons cited in the FGDs for the declining seriousness of schistosomiasis included its curability and the fact that fewer people talk about it today. Despite this, schistosomiasis remains a serious disease that reinforces poverty. As recent reports suggest, the disease exacerbates malnutrition, stunts physical and cognitive development, and reduces educational attainment, labor supply, and incomes – all of which hinder economic growth and the improvement of sanitation, access to energy, water, education, and healthcare [43].

### Optimism meets obstacles: positive attitudes hindered by limited healthcare access

The high acceptability of treatment for schistosomiasis in Chiredzi is an encouraging sign, with over 97% of respondents expressing the need for preventative measures, awareness of their infection status, and willingness to seek treatment if infected. The majority also believed that schistosomiasis could be cured (HVE: 94%, Chipimbi: 98%). Similar high levels of treatment acceptance have been reported in Shamva, Zimbabwe [15] and other regions such as Mozambique (90%) [24] and the DRC (95%) [22]. However, contrasting attitudes have been observed elsewhere, such as in Tanzania, where treatment is often rejected due to community fears and conspiracy theories about population reduction and covert birth control agendas [44].

Only a small percentage of respondents in both communities (less than 16%) considered seeking help from traditional health practitioners or religious leaders if infected with schistosomiasis. This distrust of traditional healers, particularly noted in the FGDs in Chipimbi, echoes findings from northern Senegal, where the majority of people preferred to seek treatment at local health centers [45]. Similarly, Mbereko et al. [25] observed that participants in Zimbabwe and South Africa placed greater trust in Western medicine than in traditional practices, a sentiment likely influenced by the region’s widespread Christian beliefs, which often discourage the use of traditional medicine [46].

Despite these positive attitudes, several obstacles to accessing healthcare were highlighted in the FGDs and key informant interviews. In HVE, only permanent employees of Tongaat Hulett and their families have access to the company’s private medical center, leaving temporary workers and others to seek care at distant public hospitals, which often require them to gather funds for treatment. Although treating all HVE residents may be financially challenging, extending medical services to a broader population could be beneficial, given the risk of untreated individuals reinfecting others.

In Chipimbi, healthcare access is further complicated by under-equipped facilities and a lack of medical and human resources, sometimes resulting in patients going untreated. The community currently lacks a functional clinic, with residents forced to travel up to 20 km for care, similar to the challenges faced in rural Uganda [47]. Because of this, village health workers (VHWs) serve as primary caregivers but are limited by the complexity of conditions they can treat and the availability of proper medications. The VHW program, initiated in the 1980s, plays a crucial role in disease prevention and primary care in rural and peri-urban areas and has recorded many successes in healthcare programs [30]. However, economic and health crises over the past decade have weakened community health systems, highlighting the need for regular training and support for these key personnel, whose impact extends beyond schistosomiasis control to the broader health of the community. Strengthening the VHW program aligns with the goals of the Ministry of Health and Child Care and supports efforts to achieve the 2030 NTDs elimination goals.

A significant difference was noted between the communities, with 65% of respondents in Chipimbi recognizing treatment for all infected individuals as a preventive measure against schistosomiasis, compared to only 37% in HVE. Additionally, 72.5% of HVE respondents took proactive measures to prevent bilharzia, versus 55.7% in Chipimbi, where a lack of knowledge on preventive measures was reported. This suggests the positive impact of WASH interventions in HVE. Furthermore, a substantial percentage of HVE respondents (77.5%) had received praziquantel before, compared to 54.1% in Chipimbi. The lower rates in Chipimbi were attributed to a lack of awareness or absence of treatment campaigns, which were not mentioned in any of the FGDs with Chipimbi participants. Both communities reported that school-aged children received praziquantel as part of the national control program between 2012 and 2017, though no data were available for later years.

This scarcity of mass drug administration (MDA) campaigns, particularly in rural communities like Chipimbi that relies on government input for such interventions, presents a significant obstacle to schistosomiasis control and prevention. Although Zimbabwe has conducted extensive mapping and six MDA exercises for schistosomiasis and other NTDs resulting in a great reduction of the two conditions as well as shrinking of the prevalence map as published in the National Health Strategy [30], recent reports indicate a slowdown in progress, with no one targeted or treated with MDA in 2020 [48]. This raises concerns about potentially reversing the gains so far made in controlling the disease.

### Access to safe, treated water as a determinant of risky practices

A moderate yet significant association was found between the community of residence and the practice scores of respondents (φ = 0.385, p < 0.001), implying that the community of residence influences how respondents engage in practices that minimize schistosomiasis transmission. A comparative analysis shows that more residents in HVE (64%) had moderate to good practices, compared to only 34% in Chipimbi. This difference can be attributed to the higher income levels in HVE, as a small but positive association was found between monthly income and safer practices regarding schistosomiasis risk (γ = 0.238, p = 0.001). Higher income appears to correlate with better practices [49] with possible reduction of schistosomiasis risk.

Households in HVE benefit from reliable access to tap water treated to WHO standards [50], with 98% of respondents identifying piped water as their primary source. Only a small fraction (6%) reported using natural sources like rivers, streams, or irrigation canals. Despite the general availability of taps, FGDs in HVE revealed issues with inconsistent water supply, often due to power cuts that affect water pumping, leading to the rapid depletion of stored water in tanks. Additionally, the irrigation activities at HVE significantly increase the risk of water contact, putting residents at a continued risk of schistosomiasis. The environmental health officer of HVE highlighted that the Tongaat Hulett organization, which manages HVE, is fully aware of these risks associated with intensive water use. Consequently, a fortnightly program has been implemented to inspect and treat all water bodies to control the vector, with many canals cement-lined to prevent snail habitation further reducing the potential for schistosomiasis transmission.

In contrast, Chipimbi’s situation is marked by poverty, which fosters schistosomiasis transmission as residents often have no choice but to use contaminated water for daily activities. The primary sources of water in Chipimbi are boreholes (97%), but with only seven functional ones across ten villages, natural sources like rivers, streams, and dams become essential alternatives. These sources are used for personal activities like bathing and laundry (100%) and economic activities like agriculture (84%), animal watering (54%), and fishing (54%), with 56% of respondents visiting these unsafe water sources at least twice a day. Although 86% of respondents understood the importance of avoiding water contact, they are left with limited options. FGDs in Chipimbi highlighted that while borehole water is available, its scarcity means it is reserved mainly for drinking, while river water is used for other daily activities such as laundry and bathing.

Unlike HVE, where NTDs and WASH services are managed privately by an organization, Chipimbi and other rural areas in Zimbabwe rely on the government, where coordination is more active at the national level than at subnational levels (province, district, and ward) [30]. This centralized approach may lead to delays in providing essential services, especially in marginalized communities like Chipimbi, potentially compromising the government’s goal of “leaving no one and no place behind.” The Presidential Rural Development Programme aims to address this by having the Zimbabwe National Water Authority (ZINWA) drill a borehole in each village, install reservoirs, and provide community water points with laundry facilities and model ablution facilities [51]. So far, 2,890 boreholes have been drilled under this program, with a target of 35,000. The recently launched national NTD masterplan acknowledges the government’s efforts to meet international health commitments but also highlights the insufficient funding for the NTD program, largely due to low public revenue and macroeconomic instability [30]. As a result, donor and bilateral support systems are crucial for health financing, especially for priority diseases including NTDs. This is examplified by one of the boreholes in Chipimbi being donated by China Aid. If government goals are realized, many water-borne diseases, including schistosomiasis, could be eliminated in Zimbabwe.

### No toilets, more problems!

The use of toilets is crucial in breaking the transmission cycle of schistosomiasis [52]. Nearly all respondents in both HVE and Chipimbi recognized the importance of using toilets for their health, with 98% in each community agreeing on the necessity of proper sanitation. This positive attitude contrasts sharply with findings in the DRC, where only 59% of participants shared this sentiment [22].

In HVE, toilets are widely available, with most households having either private facilities (e.g., in Chishamiso) or shared communal toilets (e.g., Section 5). Almost all respondents, except two, reported having access to a toilet, with 92% using a flush system. However, 51% admitted to occasionally defecating or urinating outside the toilet, citing reasons such as the urgency of nature’s call (53%), lack of access to toilets (26%), and insufficient water for flushing (16%). These findings suggest that merely providing toilets is not enough; regular community education on the importance of consistent toilet use is essential. FGDs revealed that this issue is particularly acute in Section 5, where shared communal toilets often become unsanitary due to water shortages, leading residents to resort to open defecation.

The situation in Chipimbi is markedly different, with a significant scarcity of toilet facilities. While 48% of respondents used pit latrines and 26% used improved pit latrines, 26% reported having no toilet facilities at all. A considerable 62% of respondents admitted to defecating or urinating outside, citing reasons such as the urgency of nature’s call (66%) and lack of access to toilets (50%). FGDs and KIIs in Chipimbi further highlighted that the scarcity of toilets was understated in the questionnaires, revealing a more dire situation. For example, in Village 1, there was an imbalanced ratio of nine toilets to 63 households (14.3%). This figure is even lower than reported by the Zimbabwe National Statistical Agency (ZIMSTAT), which found that at least 70% of households in some parts of the country lack access to toilets [53]. This falls far short of WHO recommendations for each household to have access to a toilet to curb the spread of neglected tropical diseases [54].

Promoting clean, odorless latrines is essential in rural Zimbabwe with a recent publication advocates for at least one latrine per family, ensuring the facility is kept clean and used as needed [55]. The Blair VIP latrine, developed in Zimbabwe at the Blair Research Institute (now the National Institute of Health Research) by Morgan [56], is an ideal candidate due to its simplicity, cost-effectiveness, and durability. Historical records show 100% usage of these latrines in the Mushandike Resettlement Irrigation Scheme, where they were strategically placed near fields, making them more accessible than the bush [57]. It is alarming that this toilet design is not more widespread in rural Zimbabwe several years after its local inception. This may be due to inadequate promotion by health personnel [55] and the dismantling of robust health programs due to political instability in the early 2000s, which led to the brain drain of key personnel involved in the development and advocacy of these facilities.

Toilet availability alone is not enough; proper toilet use is also critical. Despite the significant difference in toilet ownership between HVE and Chipimbi, FGDs revealed that open defecation, particularly by children, is a common issue in both areas. Children, who often see defecating in the open as a game, are the ones most infected and are contributing substantially to the transmission of schistosomiasis. Changing their behavior is essential, but this will only happen if their parents and teachers reinforce the importance of proper sanitation. However, if parents no longer view schistosomiasis as a serious disease, this behavior change will be difficult to achieve. Similar findings were reported in Uganda, where defecation behaviors were more strongly associated with infection status than household water and sanitation infrastructure [58], underscoring the need to incorporate behavior change into community-led total sanitation coverage through health and hygiene education.

### Strengths and limitations of the study

This study’s mixed-methods approach allowed for a more comprehensive understanding of the factors influencing schistosomiasis control in the communities studied. The questionnaire data indicated high levels of awareness and positive attitudes toward the disease, but it was the FGDs that provided deeper insights into these findings. For example, the FGDs revealed the connection between schistosomiasis and concerns about infertility, which helped explain the strong positive attitudes toward seeking treatment. Additionally, the FGDs uncovered the extent of open defecation practices, highlighting a significant issue that was underreported in the questionnaires – the lack of adequate and clean toilets, exacerbated by the occasional unavailability of water.

However, the study has some limitations. Notably, it did not include data on the current prevalence of schistosomiasis infection. Due to financial and permission constraints, we were unable to conduct testing within the population. This missing data limits our ability to directly correlate the observed knowledge, attitudes, and practices with actual infection rates. Although participants were asked about previous infections, this only provides a historical perspective and may not accurately reflect the current situation. Consequently, the study offers more insight into how these communities have historically managed schistosomiasis rather than providing a clear picture of their current challenges and risks.

## Conclusions and recommendations

This study reveals significant differences in knowledge, attitudes, and practices (KAP) regarding schistosomiasis between Hippo Valley Estate (HVE) and Chipimbi, two communities with distinctly different WASH (Water, Sanitation, and Hygiene) infrastructure and health interventions. Although awareness of schistosomiasis is generally high in both areas, likely due to nationwide campaigns from the 1960s and 70s, there remains a notable gap in comprehensive knowledge, particularly concerning transmission and prevention, with Chipimbi being more affected. HVE, benefiting from a long history of targeted interventions, displays better practices and attitudes toward schistosomiasis management. Residents there are more likely to engage in preventive behaviors and have a positive attitude toward medication. However, despite this, challenges in accessing treatment persist, particularly for temporary employees and residents who must rely on distant public health facilities. In contrast, Chipimbi faces severe limitations in access to water, sanitation, and healthcare facilities, leading to more risky practices and inadequate health-seeking behaviors. Despite the community’s positive attitude toward schistosomiasis treatment, the lack of available medication and proper infrastructure exacerbates the disease’s transmission. The prevalence of open defecation, despite toilet ownership, and the reliance on unsafe water sources due to inadequate boreholes further highlight the disparity between knowledge and practice. These findings underscore the critical need for tailored health education programs and significant infrastructural improvements in underserved areas like Chipimbi. The absence of clean and safe water, combined with low latrine coverage, remains a significant barrier to effective disease control. Therefore, there is an urgent need for the local government to accelerate the provision of clean water sources and promote the use of the locally designed cost-effective latrines, especially in more vulnerable regions. In conclusion, addressing the socio-ecological determinants of schistosomiasis, such as infrastructure and healthcare access, is crucial for the long-term success of control efforts in Zimbabwe and similar endemic regions. This requires a combination of improved WASH infrastructure, tailored health education, and consistent access to treatment to bridge the gap between knowledge and practice and mitigate the ongoing risk of schistosomiasis.

## Data Availability

All relevant data are within the manuscript and its Supporting Information files.

## Availability of data and materials

All data generated or analyzed during this study are included in this published article and the supplementary materials. Further inquiries or requests can be directed to the corresponding author.

## Supporting information

**Additional Table 1:** Overview of the total number of households, number of households to be interviewed, and intervals between interviewed households per section in HVE.

**Additional Text 1:** Guide for conducting semi-structured interviews with key informants chosen for their central community roles or schistosomiasis expertise

**Additional Table 2**: Sociodemographic traits of respondents.

**Additional Table 3**: Knowledge regarding schistosomiasis.

**Additional Table 4**: Full description of health-seeking practices towards schistosomiasis.

## Acknowledgements

We extend our deepest gratitude to the office of the district administration in Chiredzi for permitting this study within their jurisdiction. Our heartfelt thanks go to the community leaders, Mr. Chekenyere and Mr. Magumire of Chipimbi Community, for granting access to study areas and for their active involvement in engaging the community, ensuring local acceptance of our work. We are grateful for the valuable insights provided by Mr. White Soko of the De Beers Research Laboratory (a satellite station of the National Institute of Health Research, Ministry of Health and Child Care), Mr. Faustina Zvenyika from Hippo Valley Estates, Mrs. Otilia Chauke, Mrs. Memory Nhengeri, and Mr. Chekenyere of Chipimbi Community. Special thanks to Mr. Benny Tshuvuka of Malilangwe Trust for his crucial interpretation services to TH and RW, which significantly contributed to the success of our study. Logistical support from the Malilangwe Trust in Chiredzi was indispensable, and we thank Dr. Bruce Clegg (PhD), Mrs. Sarah Clegg, and Dr. Allan Tarugara (PhD) for their assistance. We also appreciate Dr. Tongai Mukwewa (MD) and Mr. Faustina Zvenyika for granting us access to study Hippo Valley Estates and for their invaluable insights into the local disease history. Finally, we acknowledge the dedicated local community health workers who served as community research assistants, administering the questionnaire in Hippo Valley Estates.

## Author Contributions

Conceptualization: Aspire Mudavanhu, Rachelle Weeda, Tine Huyse.

Data curation: Aspire Mudavanhu, Rachelle Weeda.

Formal analysis: Aspire Mudavanhu, Rachelle Weeda.

Funding acquisition: Aspire Mudavanhu, Rachelle Weeda, Tine Huyse.

Investigation: Aspire Mudavanhu, Rachelle Weeda, Tawanda Manyangadze Tine Huyse.

Methodology: Aspire Mudavanhu, Rachelle Weeda, Tine Huyse.

Project administration: Aspire Mudavanhu, Rachelle Weeda, Tine Huyse.

Supervision: Tine Huyse, Luc Brendock.

Writing – original draft: Aspire Mudavanhu, Rachelle Weeda.

Writing – review & editing: Aspire Mudavanhu, Rachelle Weeda, Linda Mlangeni, Tawanda Manyangadze, Tine Huyse, Maxson Kenneth Anyolitho, Luc Brendonck.

